# Identification of host endotypes using peripheral blood transcriptomics in a prospective cohort of patients with endocarditis

**DOI:** 10.1101/2023.08.08.23293811

**Authors:** Israel David Duarte-Herrera, Cecilia López-Martínez, Raquel Rodríguez-García, Diego Parra, Paula Martín-Vicente, Sara M. Exojo-Ramirez, Karla Miravete-Lagunes, Lisardo Iglesias, Marcelino González-Iglesias, Margarita Fernández-Rodríguez, Marta Carretero-Ledesma, Inés López-Alonso, Juan Gómez, Eliecer Coto, Javier Fernández, Laura Amado-Rodríguez, Guillermo M Albaiceta

## Abstract

**Objectives:** To identify endotypes in patients with infective endocarditis.

**Methods:** Thirty-two consecutive patients with infective endocarditis were studied. Clinical data and a blood sample were collected at diagnosis, and RNA sequenced. Gene expression was used to identify two clusters, termed endocarditis endotypes (EE) 1 and 2. RNA sequencing was repeated after surgery. Transcriptionally active cell populations were identified by deconvolution. Differences between endotypes in clinical data, survival, gene expression and molecular pathways involved were assessed.

**Results:** 18 and 14 patients were assigned to EE1 and EE2 respectively, with no differences in clinical data. Patients assigned to EE2 showed an enrichment in genes related to T-cell maturation and a decrease in the activation of the STAT pathway, with higher counts of active T-cells and lower counts of neutrophils. Fourteen patients (9 in EE1 and 5 in EE2) were submitted to surgery. Surgery in EE2 patients shifted gene expression towards a EE1-like profile. In-hospital mortality was higher in EE1 (56% vs 14%, p=0.027) with a hazard ratio of 12.987 (95% confidence interval 3.356 – 50] after adjustment by age and sex.

**Conclusions:** Gene expression reveals two endotypes in patients with acute endocarditis, with different underlying pathogenetic mechanisms, response to surgery and outcome.

---

Infective endocarditis is a severe disease caused by the infection of heart valves and endocardium by a pathogenic germ. Incidence of endocarditis in non-selected population is around 15-80 cases per million inhabitants and year, and increases up to 5-10 cases per 1000 inhabitants and year in high-risk groups, such as those with a prosthetic heart valve [1]. In spite of these relatively low incidences, endocarditis remains a major health issue due to its elevated mortality (in-hospital mortality of 20-30% [2,3]) and the associated resource consumption. Antimicrobial therapy and surgery remain the basis of treatment, and up to 50% of the patients require surgical replacement of the affected valves to control the infectious source and restore hemodynamics [4].

The outcome in infective endocarditis is conditioned by the interaction between the causing pathogen and the host [5]. Virulent or resistant microorganisms show higher morbidity and mortality rates. Host risk factors include previous comorbidities, history of cardiac diseases and existing intracardiac devices. Moreover, endocarditis triggers a systemic host response that may contribute to pathogenesis and outcome. The most evident cases of this exacerbated systemic response fall within the diagnosis of sepsis and are linked to the high mortality rates [6].

Recent evidence shows that critically ill patients, with a common set of symptoms and signs, may have very different underlying pathogenetic mechanisms. These specific, mechanistic-driven groups are termed endotypes [7]. The different pathogenesis may result in specific responses to therapeutic measures, that cannot be detected in trials including an unselected population. Several systemic endotypes with different immune features and mortality have been described in sepsis [8,9] and in severe infections caused by coronavirus [10].

The objective of this work is to identify the existence of endotypes in a prospective cohort of patients with infective endocarditis. Bulk RNA-seq from peripheral blood allows patient clustering according to their transcriptomic profiles at diagnosis and during their follow-up. Clinical data, outcomes and response to surgery were assessed in a cluster-specific manner, in order to identify differences in the pathogenesis that could help to find personalized treatments and, ultimately, improve the outcome in this fragile population.

## Methods

### Study design

The study protocol was reviewed and approved by Comité de Ética de la Investigación Clínica del Principado de Asturias (reference 2021.122). Informed consent was obtained from all patients or their next of kin. Inclusion criteria were an age of 18 year or more and a diagnosis of definite infective endocarditis according to Duke’s criteria [11]. Exclusion criteria were refusal to participate, immunosuppression, terminal status, or do-not-resuscitate orders.

Due to the absence of preliminary data, no formal calculation of the sample size was performed. Instead, we planned to include all patients for 1 year. All consecutive patients from April 2021 to March 2022 were thus prospectively included.

All patients followed by the hospital endocarditis team were screened. Once a diagnosis of definite endocarditis was done, informed consent was obtained, clinical data collected and a blood sample for RNA-seq drawn. Included patients were followed up to hospital discharge. In those patients in which surgery was performed, a second blood sample for RNA-seq was taken the day after the intervention. The primary endpoint was hospital death.

### RNA sequencing

Peripheral blood RNA was purified and sequenced in an Ion Torrent platform as previously described [12]. Raw fastQ files were pseudoaligned against an index (built using the GRCh38 human genome as reference) using Salmon 1.9 [13]. The resulting transcript counts were imported into R using the packages *Annotationhub* and *tximport* [14] to obtain gene counts.

### Clustering

Patients were classified into clusters at diagnosis using log2-transformed expression of the 5% genes with the largest variance. Euclidean distances were calculated, and Ward clustering algorithm applied. The two first emerging clusters were termed Endocarditis Endotypes 1 and 2 (EE1 and EE2). Clusters were represented in a two-dimension space using the UMAP algorithm.

### Analysis of differentially expressed genes

Differences in gene expression between groups of interest (either endotypes or before and after surgery) were assessed using the software package *DEseq2* for R [15]. The log2(fold change) for each gene between endotypes, with the adjusted p-value (corrected using a false discovery rate of 0.05) were calculated. Enriched pathways corresponding to genes with differential expression were identified by Gene Set Enrichment Analysis (GSEA) using the R package *clusterprofiler* [16].

### Deconvolution of cell populations

Peripheral blood gene expression was also used to identify the proportion of circulating, transcriptionally active cell populations by deconvolution using a previously validated reference matrix (*Immunostates* [17]), after removing cell types not present in peripheral blood.

### Statistical analysis

Data was collected by the research team in a dedicated database. No imputation of missing data was performed. Results are shown as median (interquartile range) or absolute count (percentage). Differences between endotypes in clinical variables or laboratory data were analyzed using a two-tailed Wilcoxon or a Chi-square test (for quantitative and qualitative data respectively). For survival assessment, a competing risks model was constructed, using death and hospital discharge alive as competing events. The hazard ratio (with its 95% confidence interval) for each outcome, after adjusting by age and sex, was calculated. All the analyses were performed using the R statistical language (version 4.2.0) with packages *ggplot2* [18]*, pheatmap* and *survival* [19], in addition to those previously cited. All the code and raw data can be found at https://github.com/Crit-Lab/endocarditis_endotypes.

## Results

Thirty-two consecutive cases of endocarditis (age 69 [62-77] years, 26 male and 6 female) were included in the study. Overall, in-hospital mortality was 37.5% (12 cases). Figure 1A shows the patient flow across the study. Table 1 shows the main demographical and clinical data at diagnosis. All patients received antibiotic treatment covering the identified germs.

**Figure 1.**
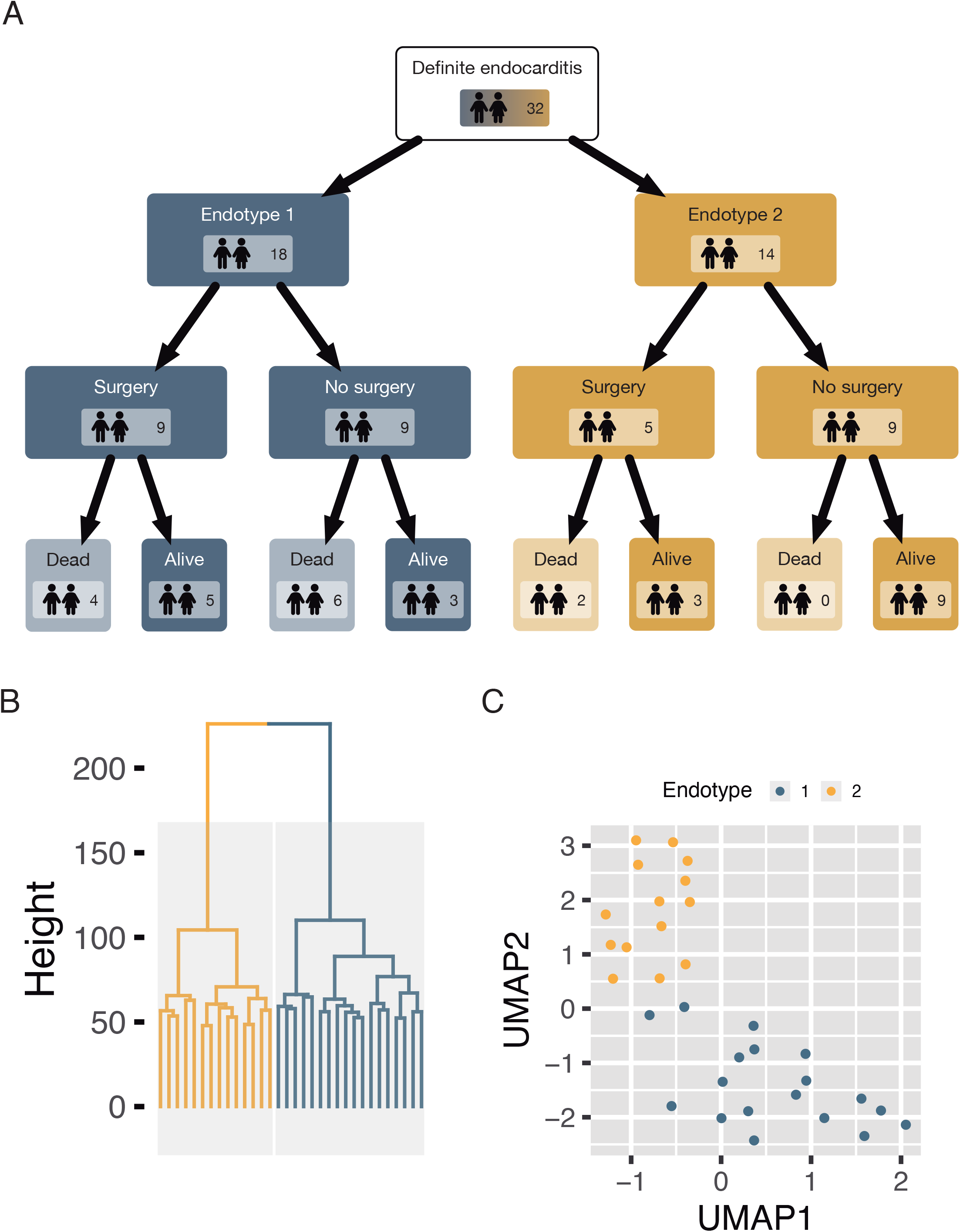
Study flow and clustering. A: Study flow diagram. B: Hierarchical clustering tree, showing the two main endocarditis endotypes (EE1/EE2). C: Uniform manifold approximation and projection (UMAP) with a bidimensional representation of each transcriptome at diagnosis.

**Table 1.**
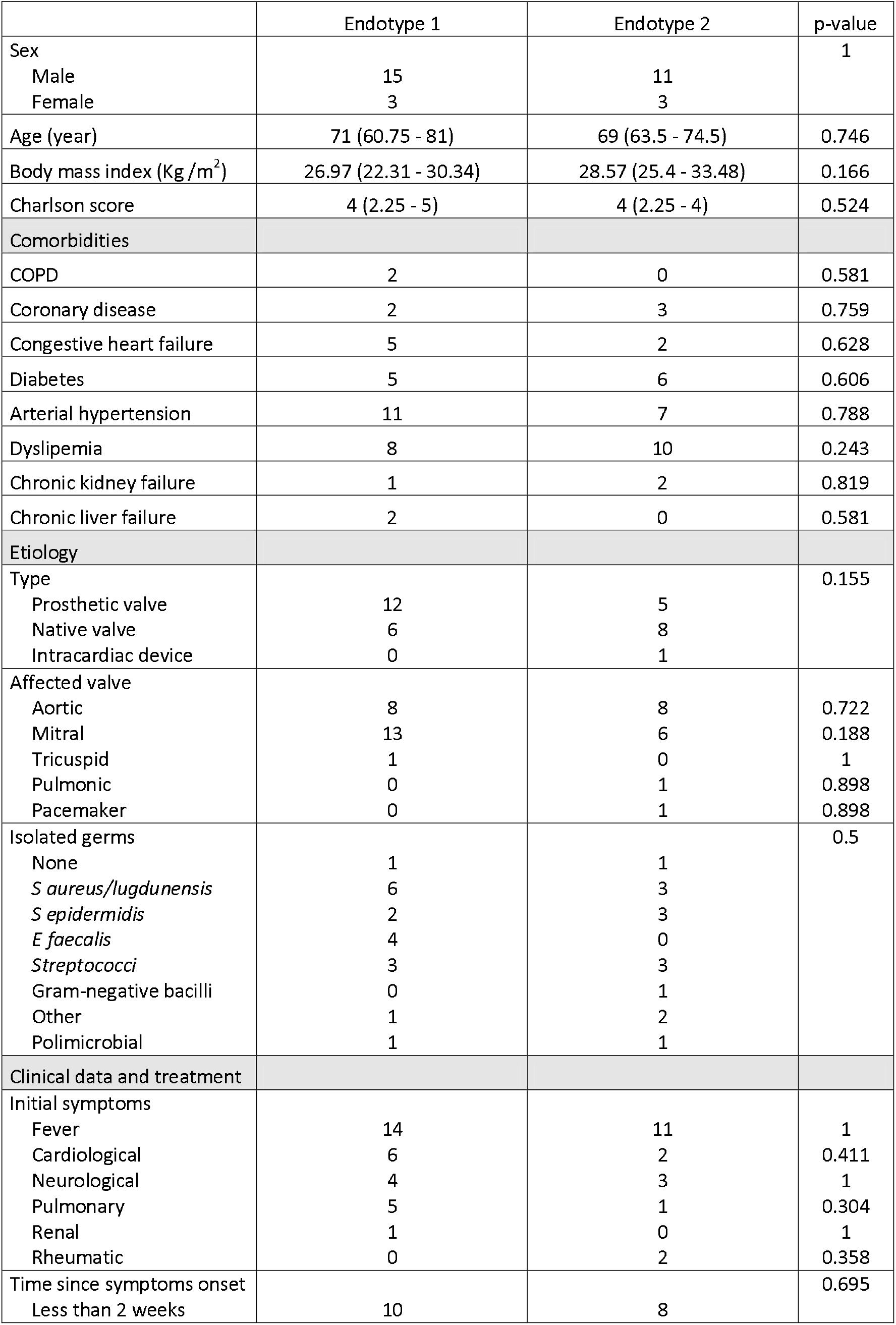
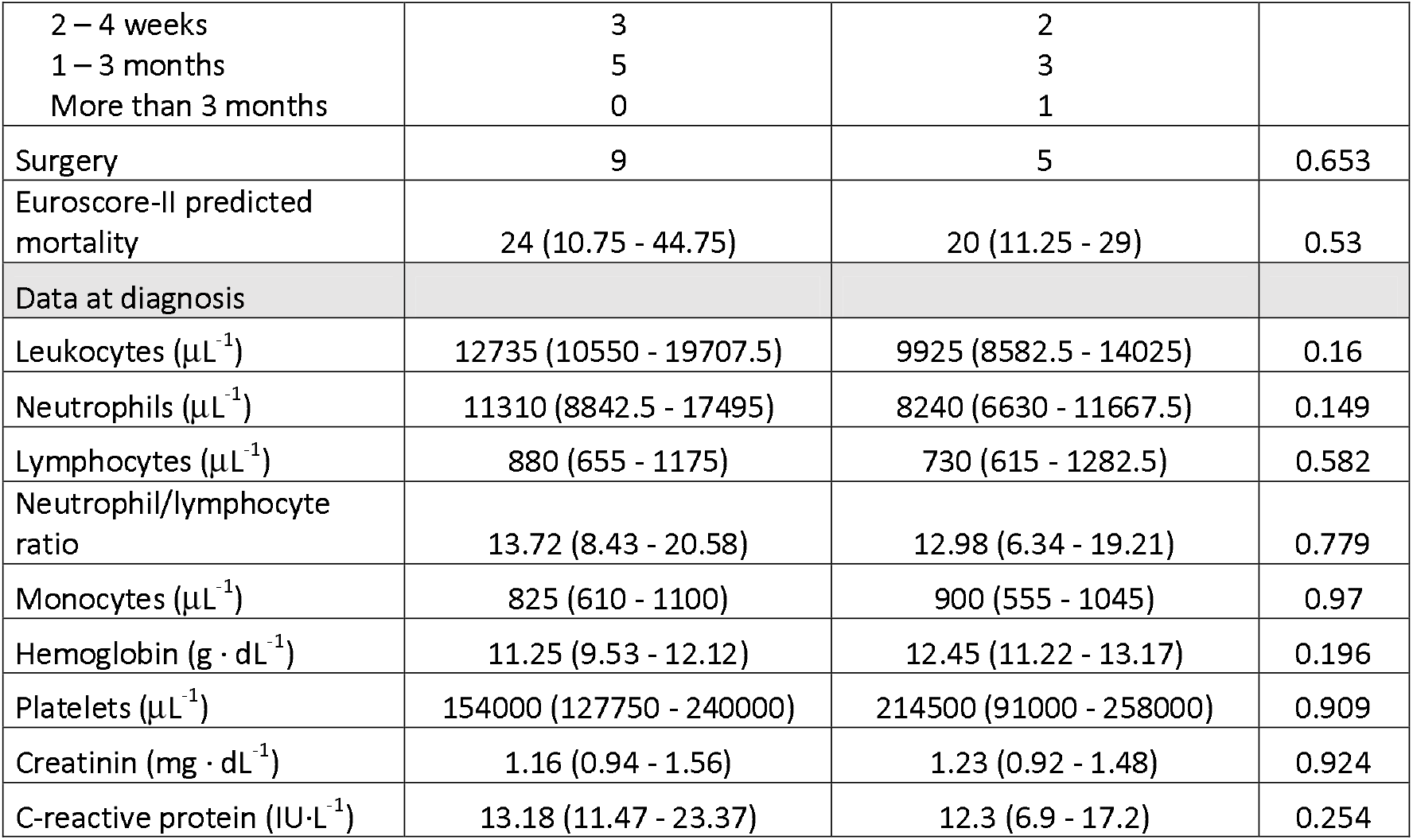
Demographical and clinical data. COPD: Chronic obstructive pulmonary disease.

### Clustering

Whole blood RNA was obtained from samples drawn at diagnosis and sequenced. Patients were clustered according to their RNA profiles. Using the 5% genes with the highest variance, two different clusters (termed endocarditis endotypes 1 and 2 [EE1 and EE2]), with 18 and 14 patients respectively, emerged (Figure 1B), with a clear separation in the UMAP projection of their transcriptomes (Figure 1C). Supplementary figure 1 shows the heatmap of these genes by cluster.

Clinical data at admission were then compared. There were no significant differences between clusters in the collected variables (Table 1).

### Differential expression analysis

There were 6577 genes with differential expression between clusters (Figure 2A). The complete list of differentially expressed genes is provided in Supplementary file 1. A Gene Set Enrichment Analysis (GSEA) was performed in these genes, revealing 199 gene ontology terms (Supplementary file 2) with an adjusted p-value lower than 0.01. Notably, several gene sets related to T cell selection and maturation were upregulated in EE2, whereas the STAT signaling pathway was downregulated (Figure 2B).

**Figure 2.**
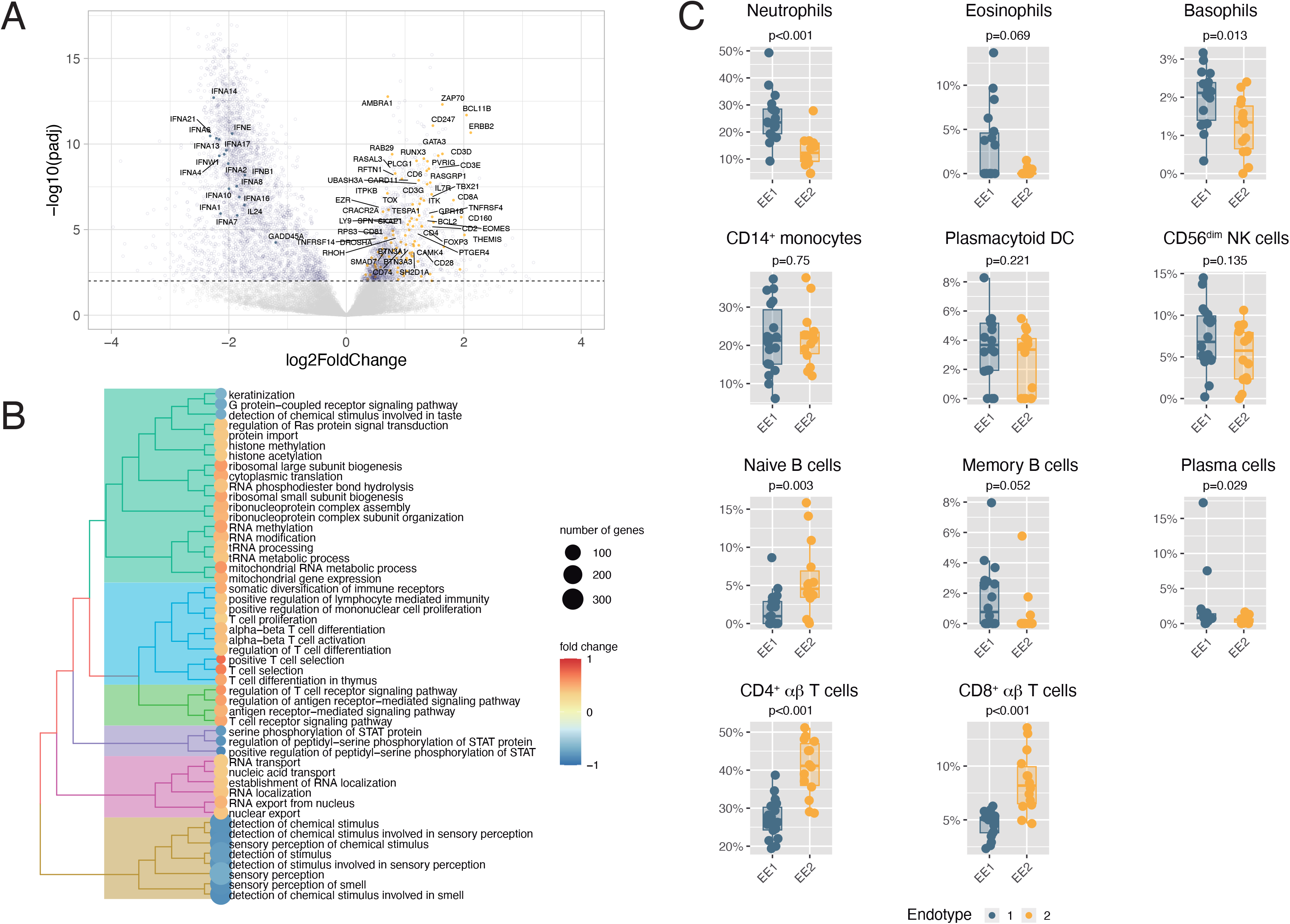
Differences in gene expression between endotypes. A: Volcano plot illustrating fold change and statistical significance for each gene. Genes corresponding to the Interferon pathway (enriched in EE1) and in T cell proliferation and differentiation (enriched in EE2) are labelled. B: Tree plot showing the pathways with differential enrichment. C: Deconvolution of transcriptionally active cell populations in peripheral blood in each endotype. P-values represent the result of a Wilcoxon test.

There were no significant differences between groups in peripheral cell counts (Table 1) or the neutrophil/lymphocyte ratio. Bulk transcriptomes were also deconvoluted to estimate the underlying transcriptionally active cell populations. Patients assigned to EE2 showed lower proportions of functional neutrophils and higher counts of T and B lymphocytes, suggesting an adaptative response to the disease (Figure 2C).

#### Differences according to microbiological results

We assessed differences in gene expression between patients with (n=18) and without (n=14) an isolation of a virulent germ (this is, Staphylococci or Enterococci). Only 1 gene (*HLA-B*) was differentially expressed. The distribution of these virulent germs was similiar across endotypes (12 in EE1 and 6 in EE2, chi-square p-value=0.323).

### Response to surgery

Nine and five patients from EE1 and EE2 respectively were submitted to surgery for valve replacement. Median time from diagnosis to surgery was 1 (0 – 4) days, with no relevant differences between clusters (1 [0-3] vs 2 [1-5], p=0.147).

In 13 of these patients (9 in EE1 and 4 in EE2 cluster), a new transcriptomic profile was obtained the day after surgery. We then compared gene expression before and after surgery in each cluster. There were 1196 and 4076 differentially expressed genes in EE1 and EE2 respectively. Among these, 795 genes with differential expression were shared between clusters. The gene-set enrichment analysis revealed that the enriched categories in EE1 were related to repression of the innate immune response and NK cell activity (Supplementary Figure 2). However, changes in EE2 after surgery resembled those observed in EE1 (in both at diagnosis and after surgery), with downregulation of pathways related to lymphocyte and T cell activation and RNA processing (Supplementary Figure 3). UMAP representation of transcriptomes before and after surgery confirms these changes in gene expression and the evolution of the profile in EE2 (Supplementary figure 4).

To find a differential response to surgery between endotypes, we identified those pathways with enrichment scores with opposite sign in each cluster after surgery (Figure 3A). First, we observed that there were no pathways with positive enriched scores, suggesting that surgery leads to a massive shutdown of gene expression in both endotypes. Differential categories included repression of neutrophil function and cytokine-mediated inflammation in EE1, and downregulation of a large variety of genetic and epigenetic mechanisms, including RNA metabolism and histone methylation in EE2. When circulating cell populations were quantified by RNA deconvolution, all the differences between clusters observed at diagnosis disappeared after surgery (Figure 3B).

**Figure 3.**
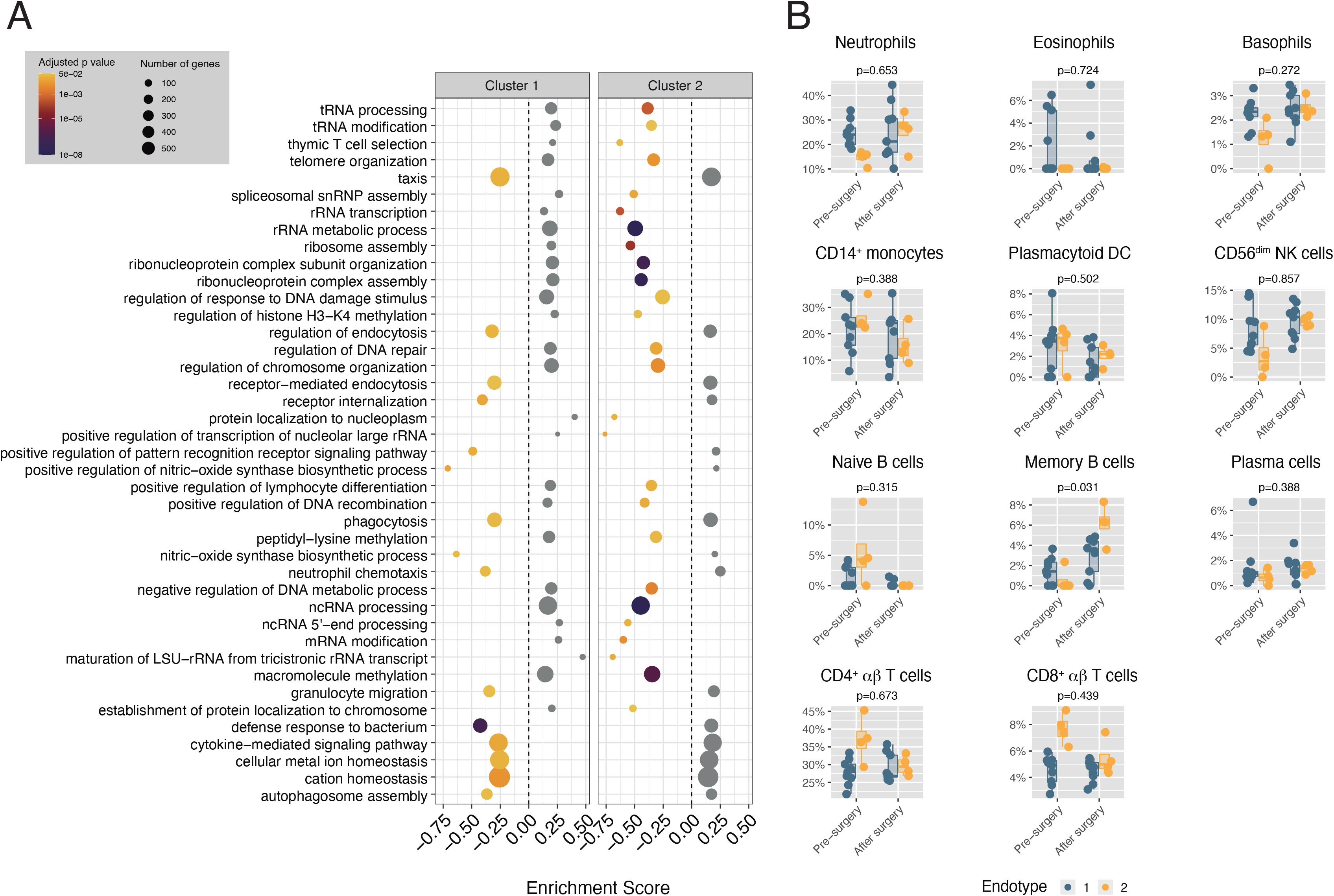
Response to surgery. A: Pathways with opposite enrichment in each endotype after surgery. B: Cell populations before and after surgery in each endotype. P-values were obtained using an analysis of the covariance (ANCOVA) to account for regression to the mean.

### Outcomes

Despite no clinical differences at diagnosis, the two identified endotypes showed different trajectories of the disease. Regarding organ failures, there were no differences in the incidence of kidney or liver failure, but patients assigned to EE1 showed a higher proportion of cardiac failure (Figure 4A).

**Figure 4.**
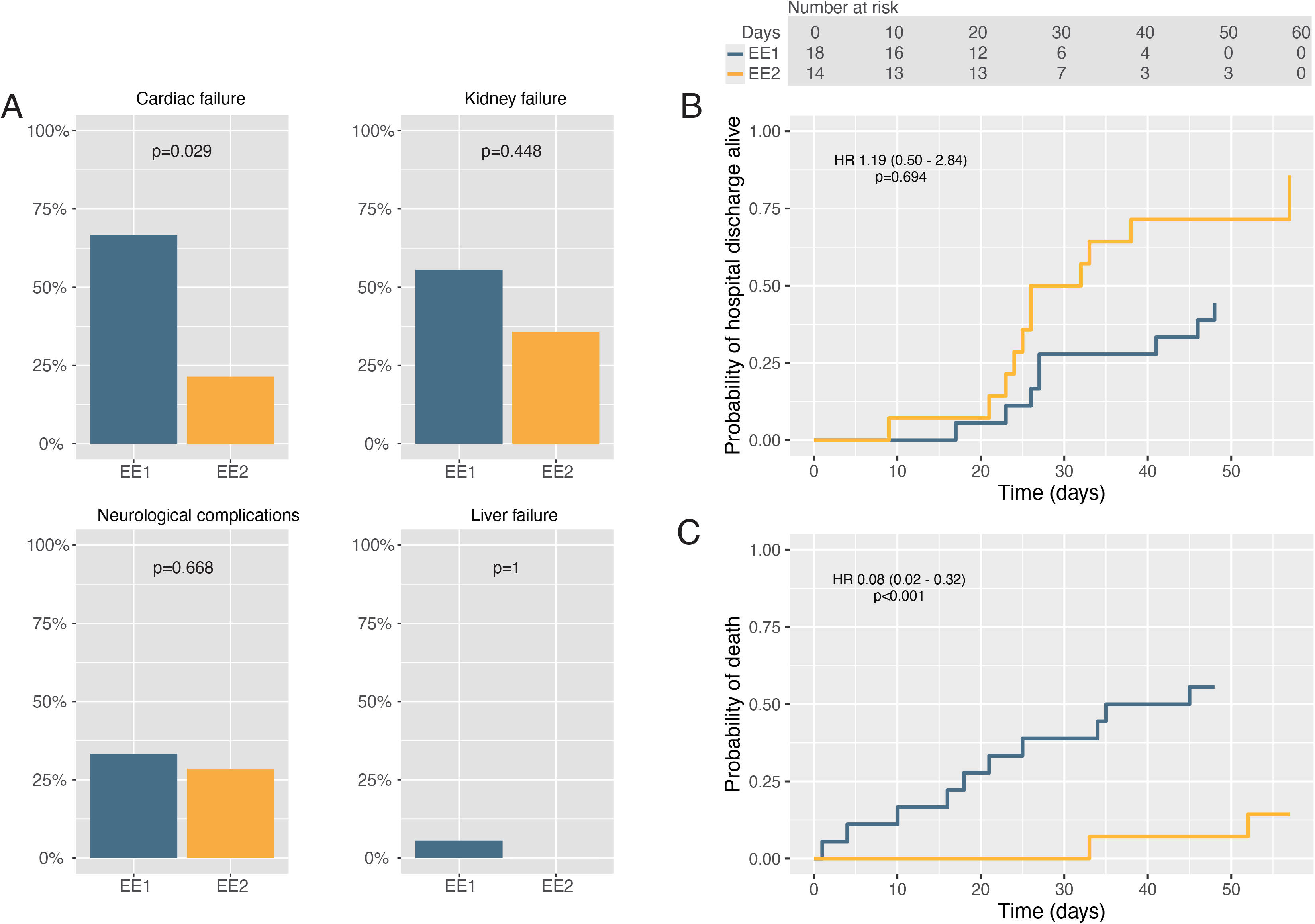
Outcomes. A: Incidence of organ failure in each endotype. B: Cumulative incidence of hospital discharge alive in each endotype. C: Cumulative incidence of death. Hazard ratios (HR) were calculated after adjustment by age and sex.

There were no differences in outcomes between patients with and without isolation of a virulent germ (Supplementary figure 5).

Patients in the EE1 cluster showed a significantly higher in-hospital mortality: 10 out of 18 patients (56%) in EE1 died, compared to 2 out of 14 (14%) in EE2 (odds ratio 7.0421 [1.075 – 83.333], Fisher’s test p-value=0.027). Amongst patients submitted to surgery, mortality was 4 out of 9 in EE1 and 2 out of 5 in EE2. In non-operated patients, mortality was 6 out of 9 in EE1 and 0 out of 9 in EE2.

When compared using a competing risks analysis, after adjusting by age and sex, assignment to EE1 was associated to higher mortality (Figure 4B), with a hazard ratio of 12.386 (3.096 – 49.554). Figures 4B and 4C show the probabilities of hospital discharge alive and death respectively.

## Discussion

Our results illustrate how transcriptomics can help to reveal two endotypes in patients with endocarditis, each one with specific pathophysiologic mechanisms, despite no major clinical differences. Moreover, response to surgery and outcome in these two endotypes is different. These findings suggest that risk prediction and therapeutic approaches in endocarditis can be personalized based on peripheral blood gene expression.

### Systemic responses in endocarditis

Transcriptomic profiling and pathway analysis allowed us to identify the underlying pathogenetic mechanisms in each endotype. EE1, linked to higher mortality rates, is characterized by the overexpression of interferon genes and the corresponding downstream activation of the STAT pathway, with an increase in functional circulating neutrophils. In opposite, EE2 patients show an increased expression of genes related to T cell activation. These changes could be interpretated as either the predominance of dysregulated or adaptative responses to the infection.

The systemic response to endocarditis depends on both pathogen- and host-related factors. Although specific bacterial strains may precipitate systemic responses due to the release of virulence factors, we did not find differences in isolated bacteria between endotypes. Other studies have identified genomic variants in interleukin-6 and interleukin-1-beta linked to a more severe systemic response to endocarditis [20]. As we did not have our patients’ genotypes, the link between genome and transcriptome cannot be clarified. However, the switch observed in EE2 patients after surgery suggests that there are environmental, non-genetic mechanisms modulating these responses [21].

Endocarditis treatment includes antibiotics and surgery to eradicate the germs, remove the source of the infection and restore valvular function and hemodynamics. Timing of surgery is still a matter of debate [4]. A randomized clinical trial [22] showed a benefit in relapse and hospitalization, but no differences at 6-month survival in patients assigned to early surgery, but the study population was younger and less severe. It is unclear how this strategy can be translated to more severe cases, in which as cardiac surgery can precipitate further deterioration [23]. Interestingly, changes in gene expression after surgery did not alter the phenotype in EE1, but caused a switch in EE2 towards a *EE1-like* profile. This raises the hypothesis that a second hit, or the accumulation of insults to the immune system, may lead to the activation of an innate, neutrophil-mediated, response that may be pathogenetic [24]. Although the reduced sample size precludes any firm conclusion, it must be noted that all the deaths in EE2 were after surgery, and maybe the delay in surgery in this group with adaptative responses can facilitate healing and avoids the accumulation of triggering events.

### Other endotypes in critically ill patients

Our findings are in line with studies identifying endotypes/subphenotypes within other critical conditions. It has been shown that septic and ARDS patients can be separated into clinically relevant subgroups. However, the identified subphenotypes are mainly driven by differences in underlying diagnoses [25]. Regarding endotypes (this is, differences in gene expression in patients with the same disease), we described two clusters in patients with severe infection caused by SARS-CoV-2 [10].Similarly, sepsis endotypes with differences in peripheral blood cell counts and pathogenesis have been described [8,9]. Notably, all these clustering analyses show that those patients in which the systemic response is characterized by neutrophilia and expression of proinflammatory cytokines show higher mortality rates, and might benefit from immunomodulatory treatments such as steroids [10]. In opposite, those patients with adaptative responses, characterized by T and B cell activation, show better outcomes and could be harmed by steroids.

### Limitations and clinical consequences

The limitations of the current study must also be highlighted. First, we performed a single measurement to categorize patients, and different clustering strategies may yield divergent results. Moreover, the sample size is reduced, and could have missed more refined endotypes. However, the identified groups have a homogeneous clinical behavior in terms of circulating cell profiles and, more importantly, outcome. An additional limitation is the lack of an external validation cohort that confirms our finding. However, we were unable to identify other published datasets with transcriptomic data in this setting. Therefore, additional studies on endotypes are warranted. Finally, treatments were not randomized, so there is risk of indication bias regarding surgery or other unmeasured confounders.

Given these limitations, our findings should be taken with caution. Current guidelines advocate for emergent surgery in endocarditis with systemic embolisms and severe hemodynamic instability [26], and are to be followed until more evidence is available. Similarly, the use of immunomodulatory agents, from steroids to specific anti-inflammatory antibodies, should only be considered within research studies.

### Conclusions

Our results show that clinically similar patients with endocarditis can be clustered in two groups with different systemic responses to both the pathogen and surgery, that result in different mortality rates. These findings may have implications to define a personalized approach to this high-risk population, in order to optimize outcome prediction and the therapeutic approach.

## Supporting information

Supplementary results

## Data Availability

Transcriptomes have been deposited at GEO (accession number GSE240321, available at https://www.ncbi.nlm.nih.gov/geo/query/acc.cgi?acc=GSE240321). All raw data and code used for analysis is available at https://github.com/Crit-Lab/endocarditis_endotypes.

https://github.com/Crit-Lab/endocarditis_endotypes

## Acknowledgements

The authors thank all the personnel at cardiac ICU from Hospital Universitario Central de Asturias for their help.

## Contributions

Conceptualization: GMA, LAR. Data Curation: RRG, IDDH, GMA. Formal analysis: IDDH, CLM, MCL, LAR, GMA. Funding acquisition: LAR, GMA. Investigation: IDDH, CLM, RRG, DP, PMV, SER, KML, LI, MGI, MFR, MCL, ILA, JG, EC, JF, LAR, GMA. Methodology: IDDH, CLM, GMA. Supervision: LAR, GMA. Writing-original draft: GMA. Writing-review and editing: All authors.

