## Supplementary results for "Identification of host endotypes using peripheral blood transcriptomics in a prospective cohort of patients with endocarditis"

**Supplementary figure 1.** Heatmap showing differences in expression between endotypes in genes used for clustering.

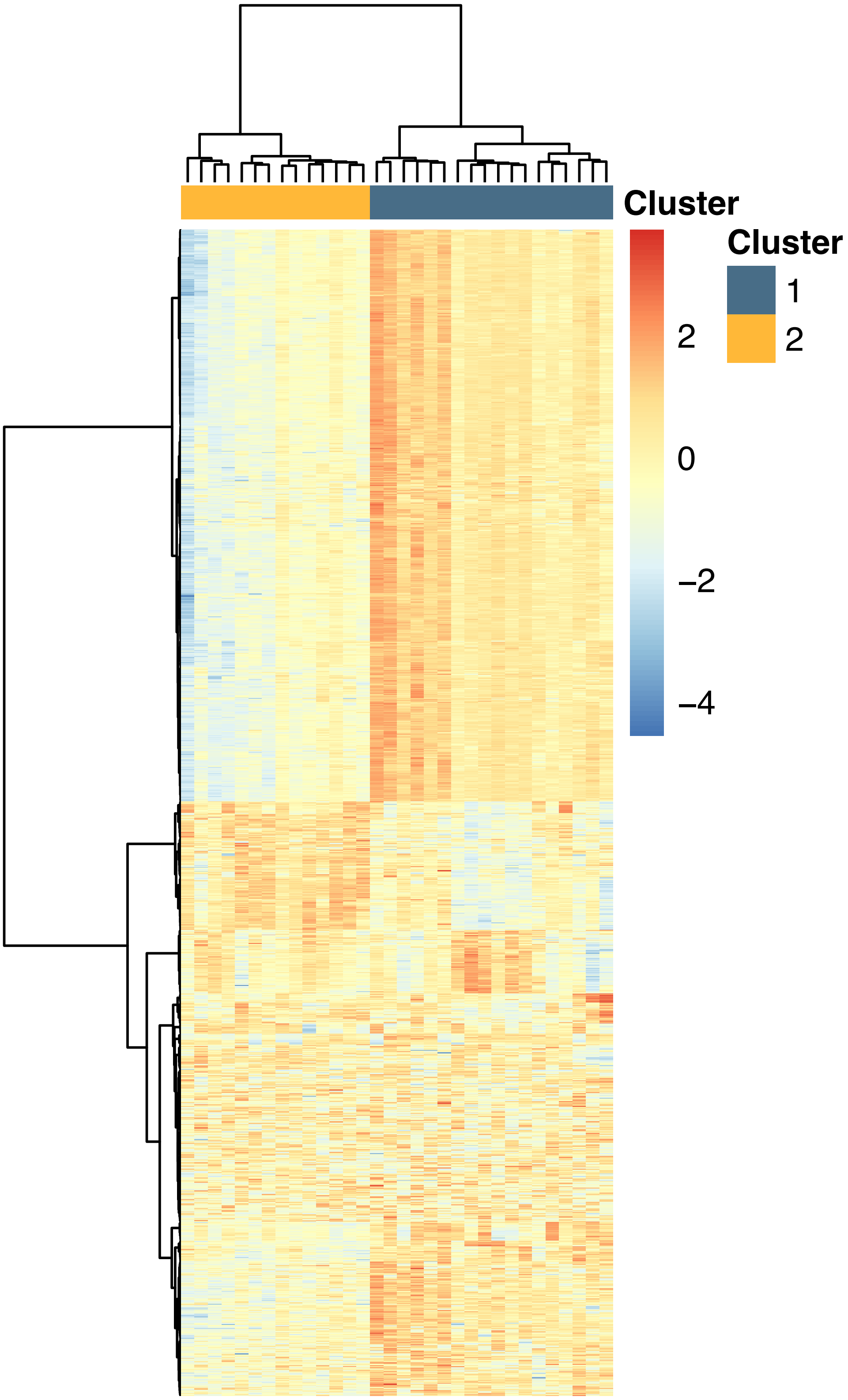

**Supplementary figure 2.** Gene ontology categories with significant changes in enrichment after surgery in patients assigned to endotype 1.

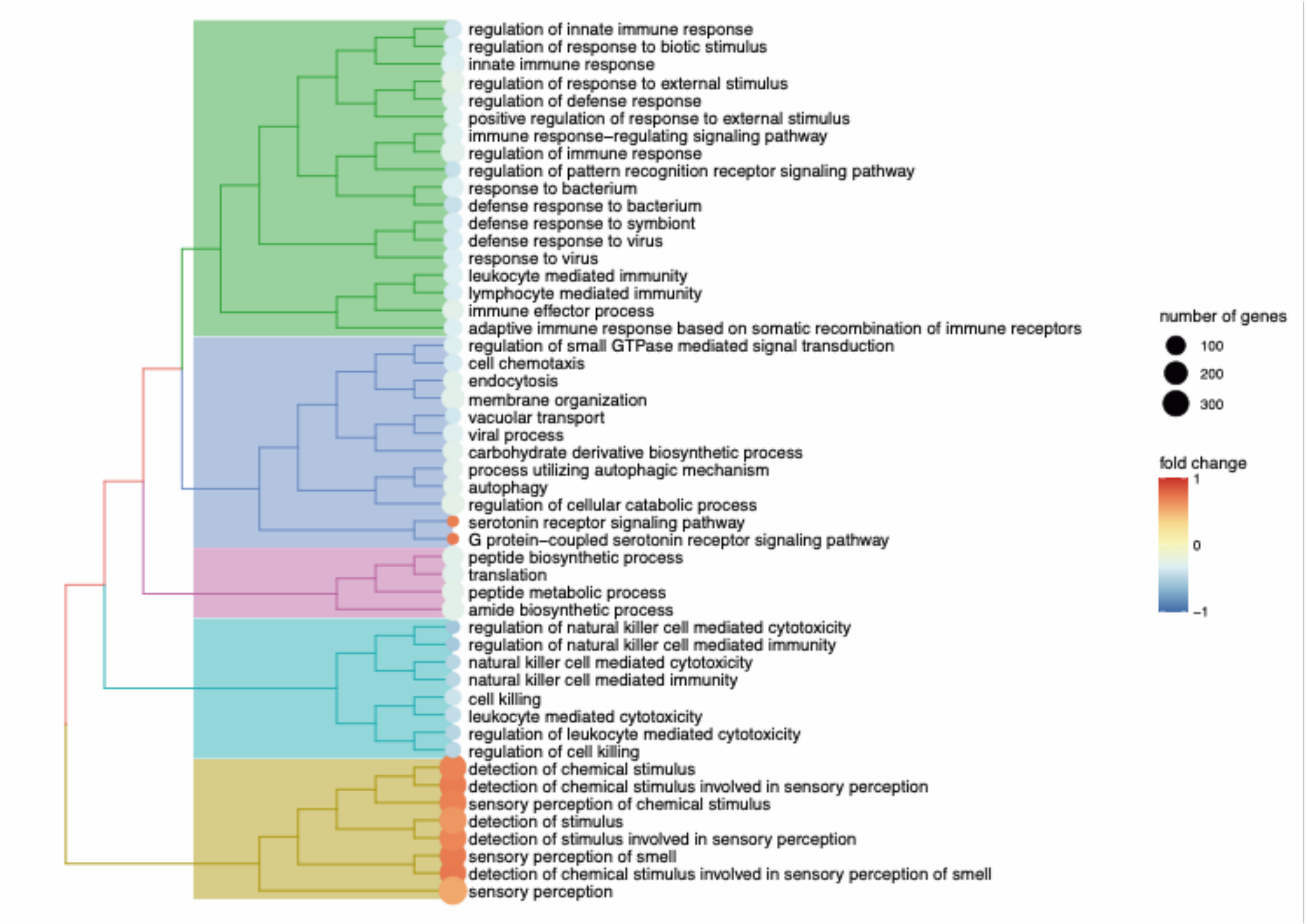

**Supplementary figure 3.** Gene ontology categories with significant changes in enrichment after surgery in patients assigned to endotype 2.

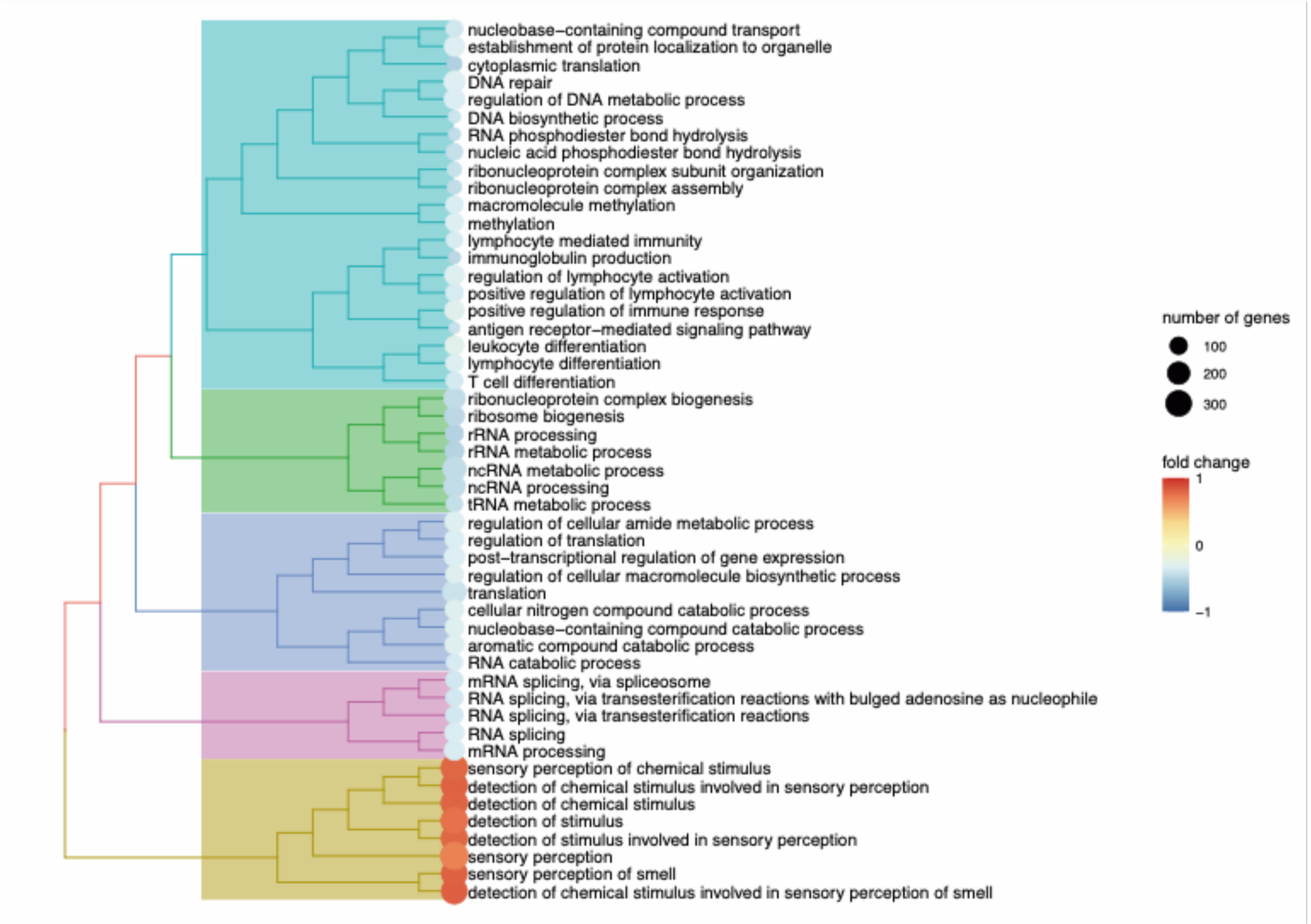

**Supplementary figure 4.** Distribution of each sample transcriptomic profile after dimensionality reduction using UMAP, categorized by endotype and timing (before and after surgery).

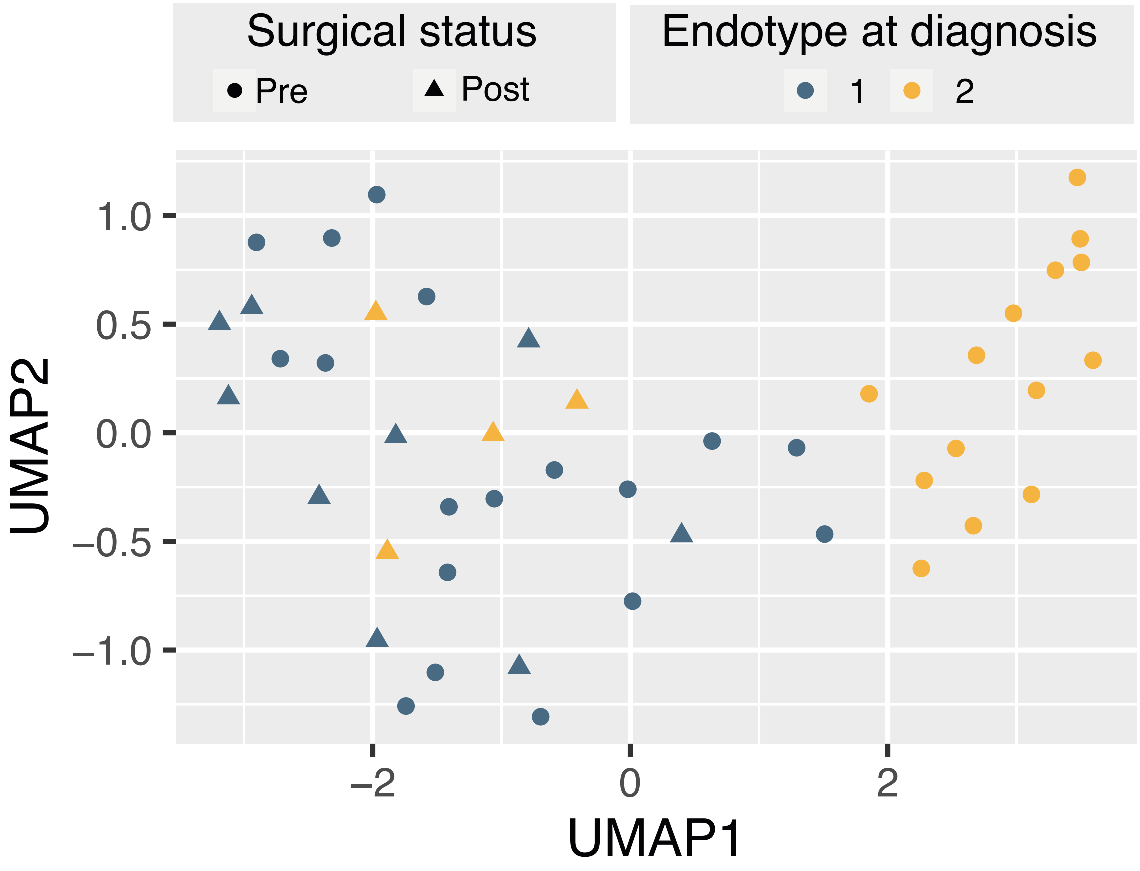

**Supplementary figure 5.** A: Cumulative incidence of hospital discharge alive in patients with and without an isolation of a virulent germ (Staphylococci o Enterococci). B: Cumulative incidence of death. Hazard ratios (HR) were calculated after adjustment by age and sex.

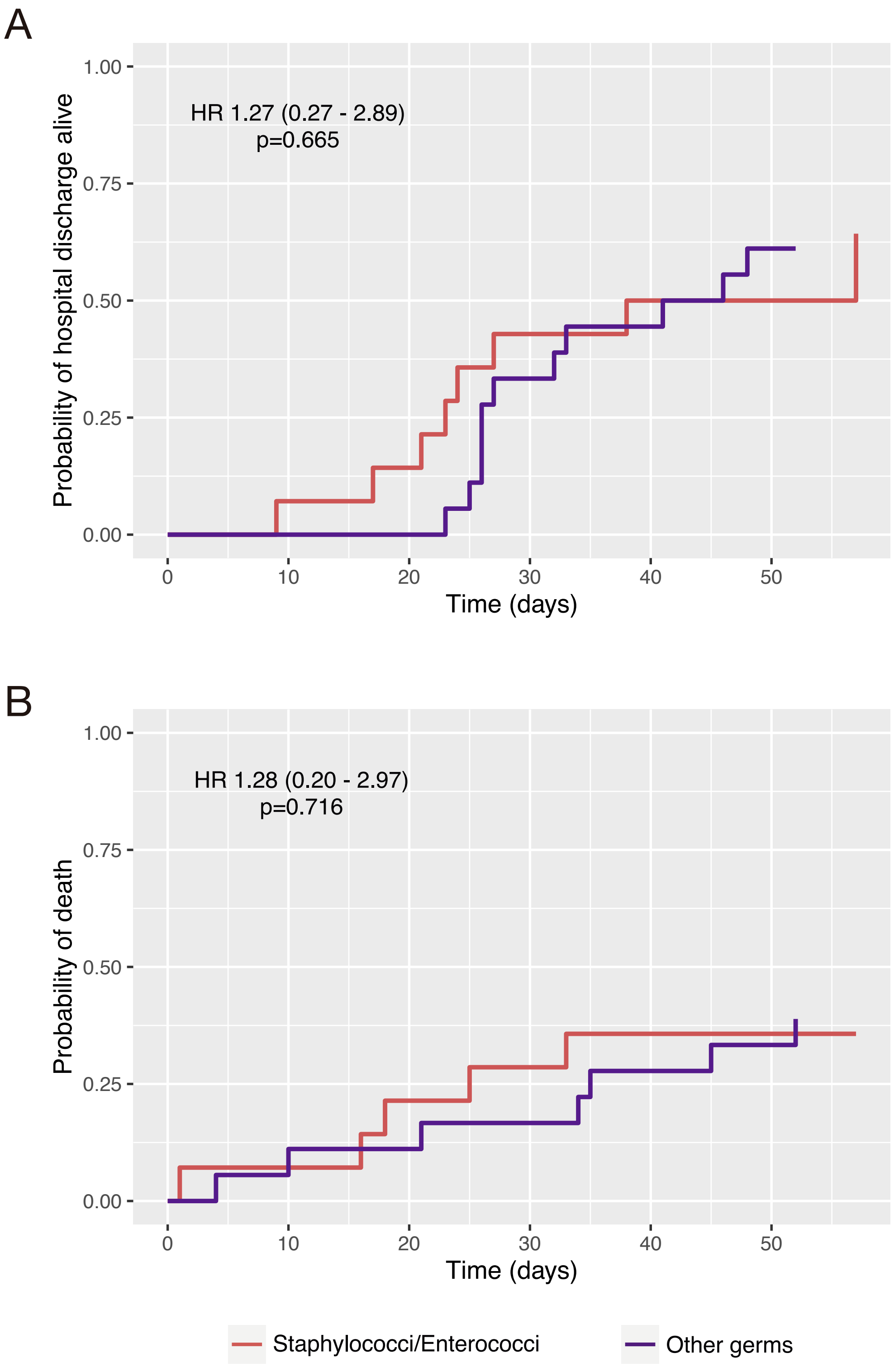
